# Quantitative and Qualitative evaluation of the recent Artificial Intelligence in Healthcare publications using Deep-Learning

**DOI:** 10.1101/2022.12.31.22284092

**Authors:** Raghav Awasthi, Shreya Mishra, Jacek B Cywinski, Kamal Maheshwari, Ashish K Khanna, Francis A. Papay, Piyush Mathur

## Abstract

**Background:** An ever-increasing number of artificial intelligence (AI) models targeting healthcare applications are developed and published every day, but their use in real-world decision-making is limited. Beyond a quantitative assessment, it is important to have a qualitative evaluation of the maturity of these publications with additional details related to trends in the type of data used type of models developed across the healthcare spectrum.

**Methods:** We assessed the maturity of selected peer-reviewed AI publications pertinent to healthcare for the years 2019–2021. For the report, the data collection was performed by PubMed search using the Boolean operators “machine learning” OR “artificial intelligence” AND “2021”, OR “2020”, OR ‘‘2019” with the English language and human subject research as of December 31, each year. All three years selected were manually classified into 26 distinct medical specialties. We used the Bidirectional Encoder Representations from Transformers (BERT) neural networks model to identify the maturity level of research publications based on their abstracts. We further classified a mature publication based on the healthcare specialty and geographical location of the article’s senior author. Finally, we manually annotated specific details from mature publications, such as model type, data type, and disease type.

**Results:** Of the 7062 publications relevant to *AI in healthcare* from 2019–2021, 385 were classified as mature. In 2019, 6.01 percent of publications were mature. 7.7 percent were mature in 2020, and 1.81 percent of publications were mature in 2021. Radiology publications had the most mature model publications across all specialties over the last three years, followed by pathology in 2019, ophthalmology in 2020, and gastroenterology in 2021. Geographical pattern analysis revealed a non-uniform distribution pattern. In 2019 and 2020, the United States ranked first with a frequency of 22 and 50, followed by China with 20 and 47. In 2021, China ranked first with 17 mature articles, followed by the United States with 11 mature articles. Imaging-based data was the primary source, and deep learning was the most frequently used modeling technique in mature publications.

**Interpretation:** Despite the growing number of publications of AI models in healthcare, only a few publications have been found to be mature with a potentially positive impact on healthcare. Globally, there is an opportunity to leverage diverse datasets and models across the health spectrum, to develop more mature models and related publications, which can fully realize the potential of AI to transform healthcare.

## Introduction

*Artificial intelligence in healthcare* is defined as the capacity of computers to mimic human cognition in the comprehension, analysis, and organization of complex medical and healthcare data^1^. AI encompasses complex algorithms that learn from the data and help in data-driven decision-making in uncertain situations. The basic objective of health-related AI applications is to examine associations between clinical procedures and patient outcomes. AI systems are used in diagnostics, treatment protocol creation, medication discovery, customized medicine, patient monitoring, care, and drug development^2^. The excitement to build artificial intelligence-based applications in healthcare is shared among clinicians, researchers, and industry^3,4^. Numerous academic departments and start-ups are building AI models to solve clinical and administrative problems. Since January 2020, numerous COVID-19-related AI models have helped in risk stratification, diagnosis, or treatment development and have been proposed for implementation in clinical care,^5^.

However, few AI models are being used in real-time for decision-making^3^. It seems imperative that researchers working in this field can robustly assess the model before deployment. Quality assessment of vast and ever-increasing AI models in healthcare is lagging^6^. In general, the quality of AI models is assessed based on predefined criteria such as Accuracy, AUROC (Area under receiver operating curve), F1-score, etc. However, it was evident that even high-performance AI models have not realized their potential after trials for real-world clinical adoption^7^. This has advocated for further validation, feasibility, and utility assessment of these AI models in clinical environments. The language of published articles, which explain the details of AI models, is the primary way to qualitatively evaluate models, which analyze their robustness and assess their maturity. The time-consuming nature of reading papers and the need to understand AI and healthcare make it difficult for humans to judge published publications. Evaluation of AI-based publications in healthcare using AI itself has recently been developed and validated^8^. This determines an answer to a maturity-level question: “Does the proposed model’s output have a direct, actionable impact on patient care by providing information to healthcare providers or automated systems?” AI-based maturity models predict the level of maturity of article^9^. In other words, maturity models, also known as ‘capability frameworks, quantitatively assess the research article. Systematic literature review and bibliometric analysis are commonly employed in all sciences to gain an in-depth understanding of a particular study subject. Recently, a BERT (Bidirectional Encoder Representation from Transformer) based language-based model was developed to assess the quality of AI models in medical literature^8^. We have attempted to evaluate peer-reviewed publications using BERT both quantitatively and also qualitatively using clinician-provided annotation in selected healthcare publications from 2019, 2020, and 2021^10,11,12^. We aimed to understand areas of healthcare that have the most mature models and what we can learn from them to advance AI in other healthcare areas. Through this evaluation framework, we have asked three key questions: 1)Maturity of *AI in healthcare* publication in various medical specialties. 2)Geographical distribution of *AI in mature healthcare* publications. 3)Distribution of various data types and model types utilized in *AI in mature healthcare* publications.

## Methods

A rigorous pipeline was employed to analyze research papers in this study **[Figure1]**. First, we utilized the recent three years of *AI in healthcare* publications ^11,10,12^ from PubMed, which had then been manually classified into 26 distinct medical specialties. We determined the nation of origin of the senior authors using the “location-tagger”^13^ python package, which employs the NER (Named Entity Recognition) NLP task. Location-tagger can detect and extract locations (countries, regions, states, and cities) from text or URLs and find relationships among countries, regions, and cities.

**Figure 1:**
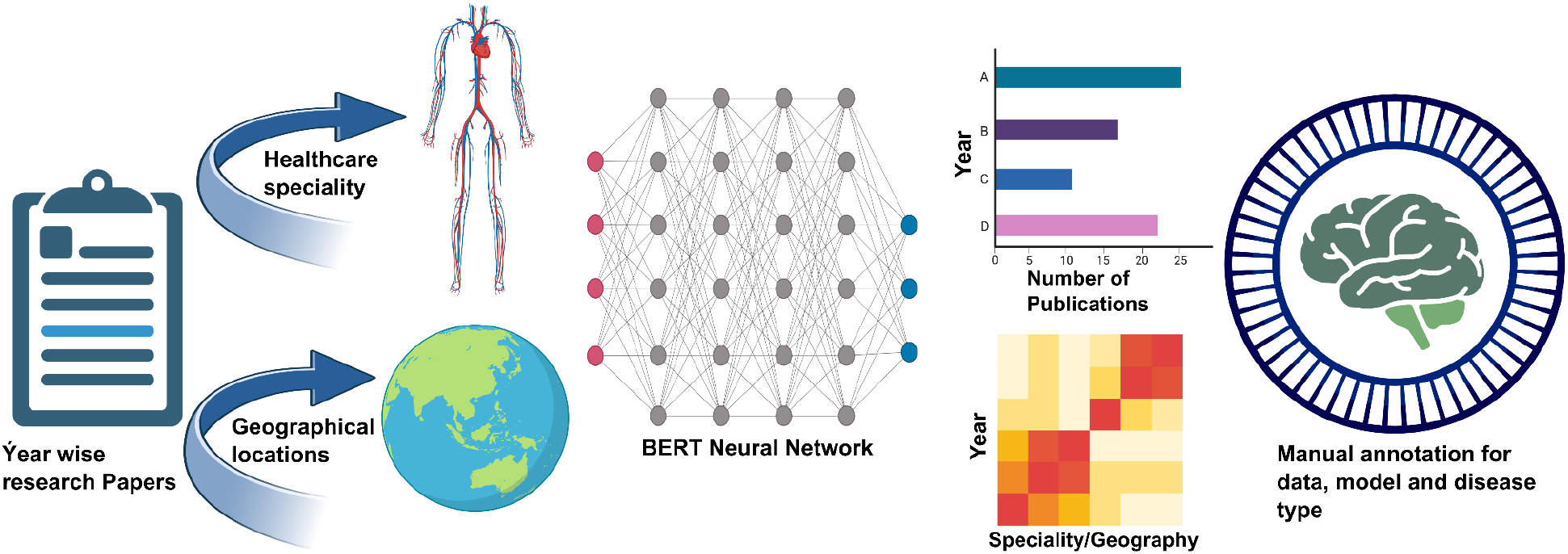
Methodology Pipeline: First, we used the most recent three years of AI in healthcare papers from PubMed, which were then manually categorized into 26 medical disciplines. We identified the nationality of senior authors. Then, we used the BERT neural networks model to determine the degree of maturity of research articles based on their abstracts. Finally, we manually annotated the mature publications with precise information, including model and data types.

Following that, we used the BERT neural networks^8^ model to identify the maturity level of research publications based on their abstracts. Finally, we manually annotated specific details from the mature articles, such as model type, data type, and disease type.

### *AI in healthcare* publication selection and data extraction

In this study, we used in-house data compiled for “Artificial Intelligence in Healthcare” reviews for 2019–2021. Data collection was performed by PubMed search using the phrases “machine learning” or “artificial intelligence” and “2021,” “2020,” and “2019” with the English language and human subject research as of December 31, each year. This search produced a preliminary list of 3351, 5885, and 4164 papers in 2019, 2020, and 2021 respectively. The papers were then individually examined and excluded based on flaws in PubMed search results or relevance to this study. Our final cohort included 1647, 3232, and 2182 papers chosen, examined, and classified into one or more medical disciplines in the years 2019, 2020, and 2021 **[Table1]**. A significant proportion of the excluded publications focused on robotic surgeries with no relevance to ML/AI, specific gene research with limited therapeutic significance, non-human investigations, or brief remarks. In each relevant specialty, 5% of articles relevant to two or more specializations were mentioned. Most drug discovery-related publications, as well as some review or editorial articles, were categorized as “General.” Using the Python geocoding module, we determined the geographical location of author connections. The location included in MEDLINE metadata refers to the country of publication and not necessarily the country where the study was undertaken. We determined the country of study based on the final corresponding author affiliation.

**Table 1.** Publications related to artificial intelligence in healthcare [Total(selected) = Publications selected after exclusions from initial PubMed search; Excluded = publications excluded based on exclusion criteria; Total (search results) = Publications based on PubMed search]^12^

| Healthcare specialty | 2019 | 2020 | 2021 |
| --- | --- | --- | --- |
| Administrative | 76 | 102 | 72 |
| Anesthesiology | 14 | 38 | 18 |
| Cardiology | 88 | 188 | 119 |
| COVID -19 | 0 | 322 | 134 |
| Critical Care | 32 | 41 | 24 |
| Dermatology | 35 | 45 | 30 |
| Education | 9 | 17 | 24 |
| Emergency Medicine | 8 | 18 | 10 |
| Endocrinology | 17 | 42 | 26 |
| Gastroenterology | 42 | 81 | 173 |
| General | 343 | 510 | 451 |
| Genetics | 114 | 120 | 65 |
| Head & Neck | 21 | 73 | 51 |
| Nephrology | 16 | 28 | 14 |
| Neurology | 70 | 172 | 92 |
| Ob/Gyn | 22 | 38 | 19 |
| Oncology | 106 | 219 | 214 |
| Ophthalmology | 56 | 132 | 82 |
| Orthopedics/Rheumatology | 20 | 48 | 24 |
| Pathology | 77 | 105 | 71 |
| Pediatrics | 31 | 39 | 25 |
| Rehabilitation Medicine | 17 | 41 | 14 |
| Psychiatry | 65 | 101 | 74 |
| Pulmonary | 19 | 38 | 21 |
| Radiology | 400 | 657 | 318 |
| Surgery | 47 | 141 | 84 |
| Total (selected) | 1647 | 3232 | 2182 |
| Excluded | 1704 | 2653 | 1982 |
| Total (search results) | 3351 | 5885 | 4164 |

### Maturity Model

We utilized an approach^8^ developed to classify the research paper’s maturity based on its abstract. The title and abstract were utilized as a predictor of the paper’s level of maturity. 2500 manually labeled abstracts from 1998 to 2020 were utilized to fine-tune hyperparameters of the BERT PubMed classifier. BERT is a deep learning model for NLP tasks that are built on transformers. BERT’s functioning completely depends on attentional mechanisms that understand the contextual relationships between words in a text. The maturity classifier was validated on a test set (n=784) and prospectively on abstracts from 2021 (n=2494). The test set model had an accuracy of 99 percent and a precision F1 score of 93%, while the prospective validation model had an accuracy of 99 % and an F1 score of 91%. Lastly, when contrasted to curated publications from a systematic review of AI versus Clinicians^14^, we have asserted that this maturity model uses joint abstract and title of an article to forecast the paper’s maturity.

### Analysis

Using the model described above, we predicted the maturity of publications for the years 2019, 2020, and 2021 and conducted temporal analysis in the following way.

- First, we have predicted a general pattern of research maturity from 2019 to 2021.
- Second, we conducted a pattern analysis by healthcare specialty for 2019, 2020, and 2021.
- Next, We examined the pattern of *AI in mature healthcare* articles through a global lens.
- Finally, we have manually annotated the data type and model type for mature papers in 2019, 2020, and 2021.

## Results

### Maturity patterns by the year

103 (99 mature models, four systematic reviews) of the total 1647 publications published in 2019 were considered mature. In 2020, there were 3232 publications and 253 (250 mature models, 3 systematic reviews) that were deemed mature. In 2021, however, there was 2182 publications total, and only 83 (36 mature models, 47 systematic reviews) were considered mature **[Figure 2 (B)]**. Percentage level estimations indicated non-monotonic patterns in the maturation tendencies of publications. We categorized 6.01 percent of publications as mature in 2019, 7.7 percent of publications as mature in 2020, and 1.81 percent of publications as mature in 2021. Since systematic reviews do not provide concise information regarding the type of AI models and Data use; hence, they were excluded from further analysis.

**Figure 2:**
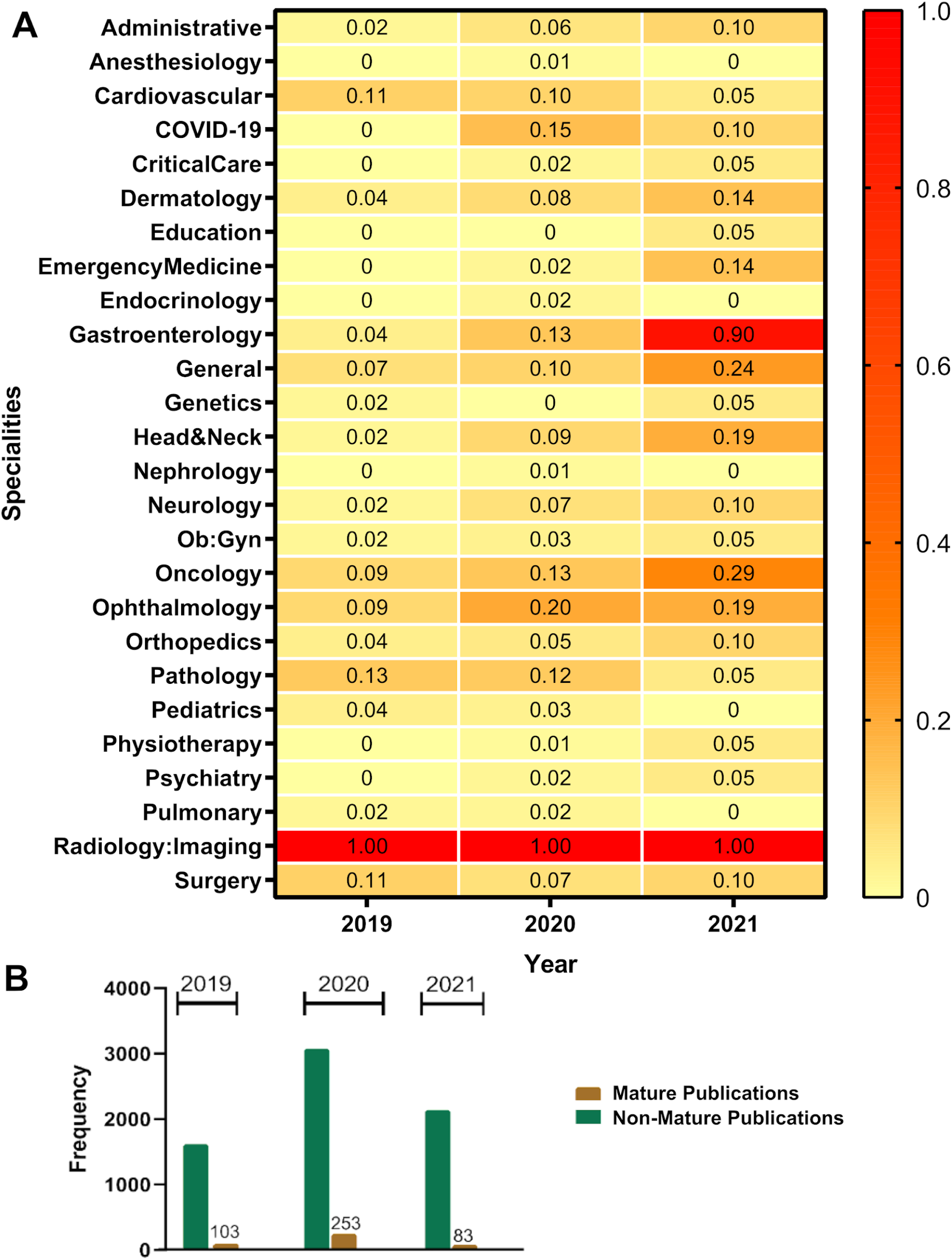
**A) Year wise pattern of mature publication by healthcare specialty:** Normalized heat map (where 0 represents the lowest number of mature publications and 1 represents the highest number of mature publications) depicts 2019–2021, ranked medical specialty in mature research publication. After classifying all of the chosen PubMed articles into one of 26 distinct medical subspecialties, we identified the overall pattern. Radiology was ranked number one in all three years. **B) Overall maturity patterns by year:** Bar graphs comparing the quantity of mature and immature publications published in 2019, 2020, and 2021. In 2019, we determined that 6.01 percent of publications were mature; in 2020, we determined that 7.7 percent of publications were mature; and in 2021, we determined that 1.81 percent of publications were mature.

### Maturity patterns by medical specialty

Different medical specialties pose unique challenges. Here, we have separated the specialty-specific findings for all 26 specialties **[Figure 2 (A)]**. Radiology has the most mature models across all specialties over three years, followed by Pathology in 2019, Ophthalmology in 2020, and Gastroenterology in 2021. Our analysis also found that the number of mature papers in Gastroenterology, Oncology, and Ophthalmology has steadily increased from 2019 to 2021. In 2020 and 2021, the COVID-19 pandemic affected the entire world. Many researchers used AI-based models to tackle this deadly infection leading to a significant number of publications. However, our analysis reveals that only 4 and 1.6 percent of these COVID-19 related publications were mature,in 2020 and 2021 respectively. Globally, cardiovascular diseases(CVD) are the major cause of mortality. In 2019, an estimated 17,9 million individuals died from CVDs, accounting for 32% of all deaths worldwide. 85 percent of these fatalities were a result of heart attacks and strokes. In 2019, 2020, and 2021, there will be 88, 188, and 119 artificial intelligence models relevant to the prognosis and prevention of cardiovascular illnesses. However, we discovered that the ranking of mature publications in CVDs fell between 2019 and 2021 compared to other specialties.

### Mature articles frequency distribution by the geographic location of the senior authors

We retrieved the country of the paper’s senior author to investigate the variation of mature papers at the level of each country **[Figure 3**]. We discovered a non-uniform distribution pattern. In 2019 and 2020, the United States ranked first with a frequency of 22 and 50, followed by China with 20 and 47. In 2021, China ranked first with 17 mature articles, followed by the United States with 11 mature articles. This indicates that mature publications are more frequent in developed nations than in developing nations. However, our geo-map analysis revealed that developing nations like India have also published mature *AI in healthcare* articles. For example, for India, we saw that in 2019 - 2021, there were only four mature publications.

**Figure 3:**
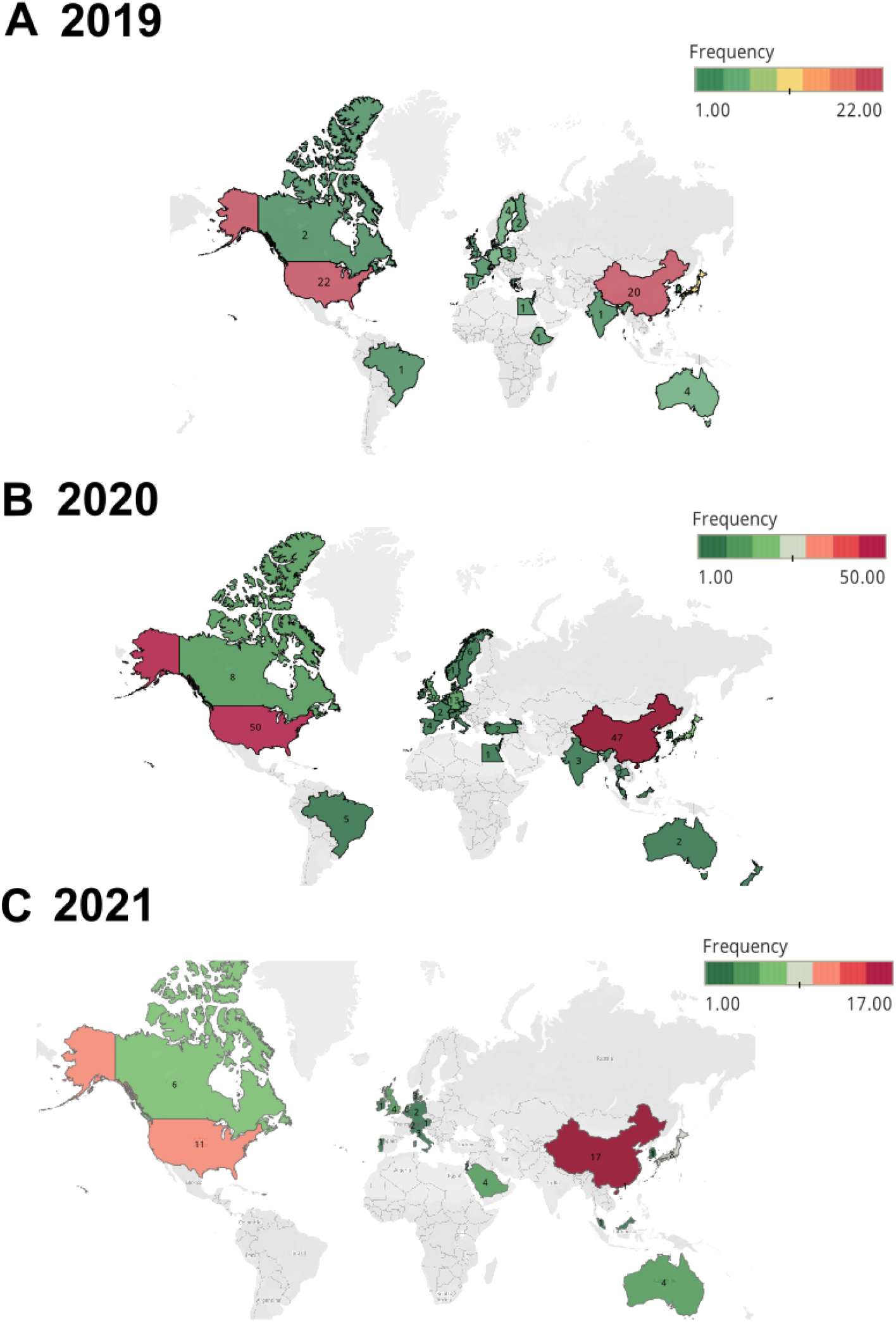
Year-wise geographical pattern of mature publication: Geo-map presents the frequency distribution of mature articles country-wise for three years. A non-uniform pattern over three years was observed. In 2019 and 2020, the highest mature publications were from the USA; in 2021, China had the highest mature publications.

### Comparison of Various Datasets and AI Models Employed in Mature Articles

We manually annotated data types and AI models within the mature articles. We have primarily categorized the data types as Image, Text, and Tabular, and model types as Deep learning (DL), Classical machine learning (ML), Natural language processing (NLP), Probabilistic models, Reinforcement learning (RL), and fundamental statistical models. Compared to textual and tabular data, we discovered that the proportion of mature publications using image data is high **[Figure 4A]**.

**Figure 4:**
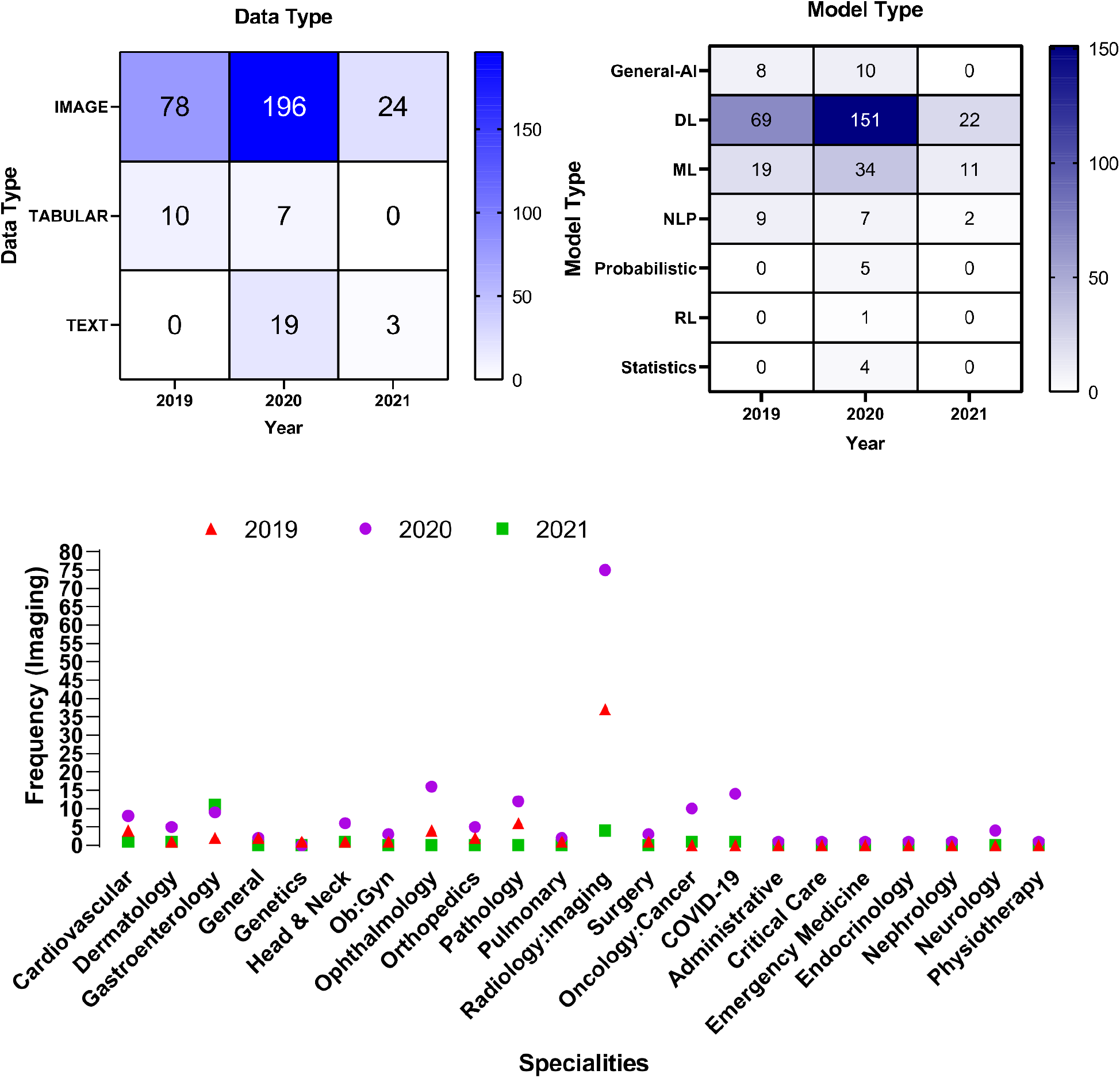
Analysis of data type and model type and disease: **A)** We subcategorized the data type into three primary categories: Image, Tabular, and Text. The heat map illustrates the mature article frequency in these three categories. The highest prevalence of Image data was recorded in each of the three years. **B)** Model type was subcategorized into frequently used model types, such as Deep Learning, Machine Learning, Natural language processing, Reinforcement learning, and Statistical modeling. The greatest proportion of mature papers using deep learning models was reported across all three years. **C)** Mature articles that used images were plotted according to their frequency of appearance in each medical specialty. It was discovered that radiology was the top among all specialties.

In 2019, 89% of mature publications incorporated image data, the same as in 2020 and 2021 (88.23% and 88.66%). From 2019 to 2020, the use of Tabular data in mature models declined from 11% to 3%, and in 2021, no mature articles used tabular data. We also discovered that text data in mature publications climbed by 8% from 2019 to 2022, with 11% of mature publications using text data in 2021. We further subdivided the use of mature publications that included image data by medical specialty. Image data were the most used in the specialty of Radiology, followed by COVID-19 as a specialty disease and Ophthalmology.

Similarly, we saw that the proportion of mature publications using Deep learning models relative to other AI models was very high **[Figure 4B]**. We observed that DL was used in 66% of all mature publications in 2019, 71% in 2020, and 62% in 2021. According to our findings, traditional machine learning models placed second behind deep learning models. 19, 34, and 11 of all mature publications used machine learning techniques in the three years examined.

## Discussion

It’s no surprise that in recent years, from 2019-2021, we identified thousands of peer-reviewed publications related to healthcare artificial intelligence (AI). However, only 5% (385/7062) of the publications were classified as mature, underscoring the urgent need for the development of clinically relevant and deployable AI models.

Although AI development in healthcare is expanding globally, according to our geographical pattern analysis, mature publications in *AI in healthcare* are concentrated in a handful of countries. The United States continues to lead in the publication of mature models, closely followed by China in year-over-year comparisons **[Figure 3]**^8^. We found some population areas, such as South America, Eastern Europe, and Africa, to be underrepresented in AI publications, which is concerning and can lead to the development of biased models and subsequently limit the generalizability and scalability of developed AI solutions^15^. For advanced AI research the availability of digitized data, healthcare information technology infrastructure, data scientists, computing capabilities, and funding are critical components which evidently are concentrated in developed countries.

To understand which specific healthcare specialties lead the AI research, we annotated the data type and speciality for the mature publication and determined that imaging data was the most prevalent. Imaging data has been the most utilized data type, probably due to easier access to open-source data supported by various institutions such as Harvard, MIT, Stanford ^16^, and the Radiological Society of North America (RSNA)^17^. Imaging data used to develop mature models included various modalities, such as computed tomography (CT scans), magnetic resonance imaging (MRI), and simple radiographs (X-rays). Early interest in adopting image-based AI for ophthalmologic disease diagnosis, such as diabetic retinopathy, has also been supported by the increased availability of fundoscopic images^18^. Imaging data in some mature models also included cine loops, particularly in specialties such as Gastroenterology (endoscopy videos) and Cardiology (echocardiography cine loops)^19^.

In 2009, Imagenet started off the revolution in the general use of image interpretation AI solutions, especially Convolutional Neural Networks (CNN)^20^. Following similar patterns, in healthcare, CNN continues to be the most commonly used AI model, particularly for the interpretation of imaging data **[Figure 4]**. The proliferation of research in the automated classification of lung nodules on chest X-rays or for stroke diagnosis has led to the further development of mature models in Radiology^21^. Similarly, the adoption of AI for diagnosing ophthalmologic diseases such as diabetic retinopathy spurred an increase in research and development from industry and healthcare entities, which continue to evolve further and mature ^22^. In specialties such as cardiology and gastroenterology, the use of deep learning in enhancing echocardiography image acquisition and interpretation or endoscopy has resulted in an increased number of publications describing mature models^23,24^. Many of these models, after FDA approval, have been embedded in medical devices or clinical workflows^7^. Unlike CNN-based models, large language models or multimodal models have been developed more recently. Publications using text data or multimodal data have been steadily increasing, and their maturity is improving^25,26^.

Readily available CNN algorithms and large imaging data repositories enabled radiology and other image-based specialties such as ophthalmology, gastroenterology, oncology, and cardiology to generate a huge growth of mature model publication. ^27^ **[Figure 2 (A)]**. Similar to radiology, AI applications from these specialties are also being implemented in healthcare. Oncology-based mature models are primarily based on imaging data with the use of deep learning algorithms ^28^ COVID - 19 presented a unique opportunity for researchers to apply some of the methods from imaging-based modeling to interpret chest X-rays and CT scans, amongst others ^29^. Although progress was made in a relatively short time to create mature models and publishing, adoption in real life has been limited, especially now that the pandemic slowed down^30^.

While many other search methodologies to evaluate more publications using publication databases such as Scopus could have been utilized, we decided to use Pubmed due to the ready availability of a validated maturity model using PubMed and related data. Conference abstracts or publications which were not in the English language might have led to some loss of data in our evaluation. Still, we believe our methodology captures most of the publications and addresses the purpose of our evaluation.Also, while there are various publication ranking methods, such as a number of citations that can be used, they have limited value in shorter evaluation time frames.

The application of AI in healthcare has caught the imagination of many, leading to an exponential rise in the number of publications over the past few years. Our evaluation demonstrates the potential and the opportunity to utilize the available data fully and diverse AI models across the world and the entire healthcare domain.

## Data Availability

We utilized open-sourced data for our study.

https://www.researchgate.net/publication/358897338_Artificial_Intelligence_in_Healthcare_2021_Year_in_Review

https://www.researchgate.net/publication/349570341_Artificial_Intelligence_in_Healthcare_2020_Year_in_Review

https://www.researchgate.net/publication/340926403_2019_YEAR_IN_REVIEW_MACHINE_LEARNING_IN_HEALTHCARE

